# Nasal CPAP is associated with increased midfacial retrusion in children with *FGFR*-related craniosynostoses: risk factors and biomechanical determinants

**DOI:** 10.64898/2026.10.21.26366177

**Authors:** Maxime Taverne, Margaux Ntirubuza, Yehong Zhong, Aloïs Roussel, Suzann Beukes, Ombline Delassus, Lucie Griffon, Mehran Moazen, Sébastien Laporte, Christophe Muth-Seng, Philippe Rouch, Giovanna Paternoster, Eric Arnaud, Corinne Collet, Brigitte Fauroux, Sonia Khirani, Roman H. Khonsari

**Affiliations:** Craniofacial Growth and Form Laboratory, Imagine Institute, INSERM UMR1163, Université Paris Cité, Paris, France; Department of Craniomaxillofacial Surgery and Digital Technology Center, Plastic Surgery Hospital, Peking Union Medical College and Chinese Academy of Medical Sciences, Beijing, China; Pediatric Noninvasive Ventilation and Sleep Unit, Hôpital Necker – Enfants malades, Assistance Publique – Hôpitaux de Paris, Paris, France; VIFASOM UMR 7330, Université Paris Cité, Paris, France; Department of Mechanical Engineering, University College London, London, United Kingdom; Institut de Biomécanique Humaine Georges Charpak (I-Biomec), Arts et Métiers ParisTech, Paris, France; Department of Pediatric Neurosurgery, Hôpital Necker – Enfants malades, Assistance Publique – Hôpitaux de Paris, Paris, France; Clinique Marcel Sembat, Ramsay Générale de Santé, Boulogne-Billancourt, France; Service de médecine génomique des maladies rares, Hôpital Necker – Enfants malades, Assistance Publique – Hôpitaux de Paris, Paris, France; ASV Santé, Gennevilliers, France; Service de chirurgie maxillofaciale et chirurgie plastique, Hôpital Necker – Enfants malades, Assistance Publique – Hôpitaux de Paris, Paris, France

## Abstract

**Background:** Children with *FGFR*-related craniosynostoses frequently require prolonged nasal continuous positive airway pressure (CPAP) for severe obstructive sleep apnea while definitive craniofacial procedures are delayed. We investigated whether sustained CPAP use alters midfacial growth and identified the morphological and biomechanical determinants of this effect.

**Methods:** In this retrospective observational study, we combined longitudinal 3D morphometric analyses of craniofacial shape in CPAP-treated and untreated children with *FGFR*-related craniosynostoses with experimental measurements of mask-induced forces and pressure distribution using 3D-printed patient-specific facial models instrumented with sensors. Multivariate analyses characterized growth trajectories, and structural equation modeling (SEM) disentangled the effects of three major risk factors: age, initial morphology, and CPAP duration.

**Results:** CPAP-treated patients exhibited a distinct growth trajectory characterized by greater midfacial retrusion than untreated patients with similar baseline morphology. Mask use generated mechanical loads of 6–10 N. Intentional overtightening markedly increased the total force applied to the face, but no notable difference was observed between expert and non-expert operators. Craniofacial morphology was the main determinant of force and pressure distributions and of growth response, identifying morphological risk factors. CPAP duration exaggerated these effects, particularly at younger ages. SEM supported links between morphology, mechanical loading, and deviation from expected growth.

**Conclusions:** Nasal CPAP is associated with additional midfacial growth deviation in children with *FGFR*-related craniosynostoses, with susceptibility strongly dependent on baseline morphology, treatment duration, and age at initiation. These findings support systematic craniofacial growth monitoring during prolonged CPAP and consideration of respiratory management and interface fit within longitudinal craniofacial treatment planning.

## Introduction

*FGFR*-related craniosynostoses combine premature fusion of cranial vault and facial sutures and skull base synchondroses, with intricate morphological and functional impairments, including brachycephaly, exorbitism, midfacial hypoplasia, and craniocervical junction anomalies. Obstructive sleep apnea (OSA) is a major early complication, affecting almost half of patients during infancy or childhood [1,2], most often because midfacial hypoplasia reduces upper airway dimensions [3]. Chronic obstruction may further impair craniofacial growth through oral breathing, potentially aggravating maxillary deficiency [4].

Management of OSA in *FGFR*-related syndromes must balance respiratory severity, craniofacial growth, and surgical timing. In infants, options include upper-airway ENT procedures, continuous positive airway pressure (CPAP), and tracheostomy. Craniofacial advancement procedures are generally reserved for older children. Midface advancement can enlarge the upper airway and improve nasal breathing [5,6], while adenotonsillectomy may be useful in selected patients [7,8], especially before frontofacial advancement [9]. Because early surgery carries morbidity and recurrence, CPAP often serves as a non-invasive bridge for persistent severe OSA in infancy [8,10].

However, prolonged CPAP may itself affect facial development. Several reports suggest that nasal CPAP in growing children can contribute to midfacial retrusion [10,11,12]. This concern is particularly relevant in *FGFR*-related craniosynostoses, where CPAP is applied to an already abnormal growth pattern and may worsen one anatomical contributor to airway obstruction. This creates a therapeutic dilemma: delaying surgery may reduce early surgical burden but prolong mechanical constraint on the developing midface, whereas earlier surgery may reduce CPAP exposure at the cost of greater operative burden and recurrence risk. Tracheostomy remains an invasive alternative.

Nasal CPAP requires an airtight seal and sustained mask-to-face contact, producing repeated loading on the nose and upper lip. External mechanical forces can modify craniofacial growth trajectories in children [13], as shown in orthopedic treatments and other contexts of growth redirection [14–17]. Experimental studies of CPAP interfaces show heterogeneous contact pressures concentrated on the nasal root and midface, and suggest that mask fit depends on craniofacial phenotype [18–20]. Yet longitudinal growth and mechanical measurements have not been integrated, and the respective roles of contact force, treatment duration, CPAP level, age at initiation, baseline morphology, and their interactions remain unclear.

We therefore hypothesized that CPAP exposure is associated with additional midfacial retrusion beyond the natural course of *FGFR*-related growth; that mask-induced force and contact pressure contribute to this growth deviation; and that specific morphotypes are at greater risk of unfavorable loading. We retrospectively compared longitudinal facial growth in infants with *FGFR*-related craniosynostoses with and without CPAP and experimentally assessed nasal CPAP mask loading on 3D-printed craniofacial infant models.

## Materials and methods

### Cohort

We reported this observational retrospective study in accordance with the STROBE guidelines [21] and the recommendations of Lederer et al. regarding causal inference studies [22] (S1 Checklist). The study included 43 participants: 28 children with syndromic craniosynostosis (Crouzon-Pfeiffer and Apert syndromes) and 15 age- and sex-matched healthy controls (flowchart of patient selection in Fig 1). Healthy controls had no craniofacial abnormalities and underwent CT for unrelated indications (e.g., trauma or infection). Patients were followed at Necker – Enfants malades Hospital (Paris, France) from 2010 onward, extracted in 2024, and classified into a CPAP-treated group (“mask group”, n = 15) and a non-treated group (“no-mask group”, n = 13). No formal sample-size calculation was performed; all eligible patients meeting the inclusion criteria during the study period were included. In the mask group, CPAP was initiated because of severe OSA and delivered using standard pediatric nasal interfaces (mainly Respireo Soft Baby, Air Liquide, or Nonny, AG Industries), with pressures ranging from 6 to 10 cmH_2_O. In the no-mask group, CPAP was not indicated in the absence of OSA, or because OSA was mild or moderate with no clinical symptoms. Routine ENT procedures were performed in both groups (93% of the mask group and 54% of the no-mask group). No patients underwent midfacial advancement surgery (frontofacial monobloc advancement, Le Fort III osteotomy) during the study period. Posterior cranial expansion had been performed in most patients in both groups (87% of the mask group and 85% of the no-mask group), reflecting standard early management [23]. Full cohort details are given in S1 Table.

**Fig 1.**
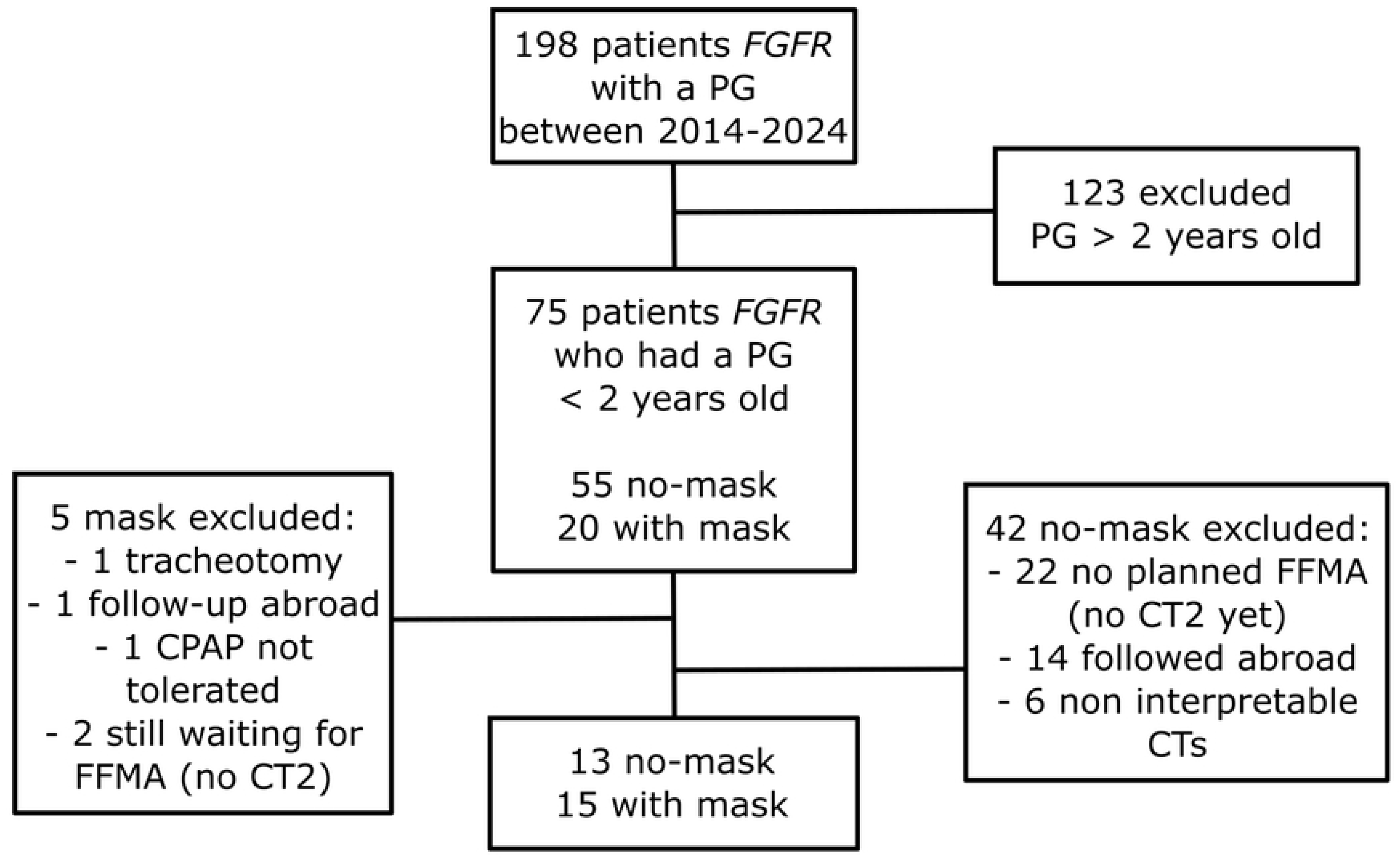
Flowchart of patient selection. PG, polysomnography; CT, computed tomography. For each patient, two CT scans were analyzed: CT1 (near CPAP initiation in the mask group) and CT2 prior to midface surgery (Fig 2.1). The time between CT1 and CT2, Δt (in days), was taken as the CPAP duration in the mask group. In the no-mask group, CT timing was matched to minimize age-related confounding effects. In healthy controls, cross-sectional CT scan timing was matched to the CT1 of patients.

### Ethics statement

This retrospective study of data collected in the context of routine care was conducted in accordance with the French reference methodology MR-004 of the Commission nationale de l’informatique et des libertés (CNIL), which governs research not involving human subjects [24]. The study was registered with the Data Protection Officer of Assistance Publique – Hôpitaux de Paris. Parents or legal guardians were informed of the study and did not object to the use of their child’s data; under this framework, written informed consent is not required. Data were pseudonymized before analysis.

### 3D longitudinal phenotypic data

#### Imaging data

Computed tomography (CT) images were collected and stored as digital imaging and communications in medicine (DICOM) files and segmented using ProPlan CMF 3.0 (Materialise, Leuven, Belgium). Metadata for comparative analysis included age at CPAP initiation and Δt.

#### Morphological descriptors

Thirty-four landmarks were placed on the midface and forehead by an experienced craniofacial surgeon (Y.Z.) (Fig 2.2, S2 Table). Intra-rater repeatability was assessed on repeated landmarking of three patients. Landmark coordinates were aligned using Procrustes superimposition without scaling, allowing analyses to preserve size-related information. Longitudinal shape change was quantified as the coordinate-wise difference between CT2 and CT1. Baseline morphology and longitudinal shape changes were compared between groups using Procrustes MANOVA and two-block partial least-squares (2b-PLS) regression, respectively. Thin-plate spline deformation was used to visualize the associated shape changes. Theoretical growth trajectories were generated by computing 2b-PLS regressions between vectors of deformation and log_10_-transformed Δt.

**Fig 2.**
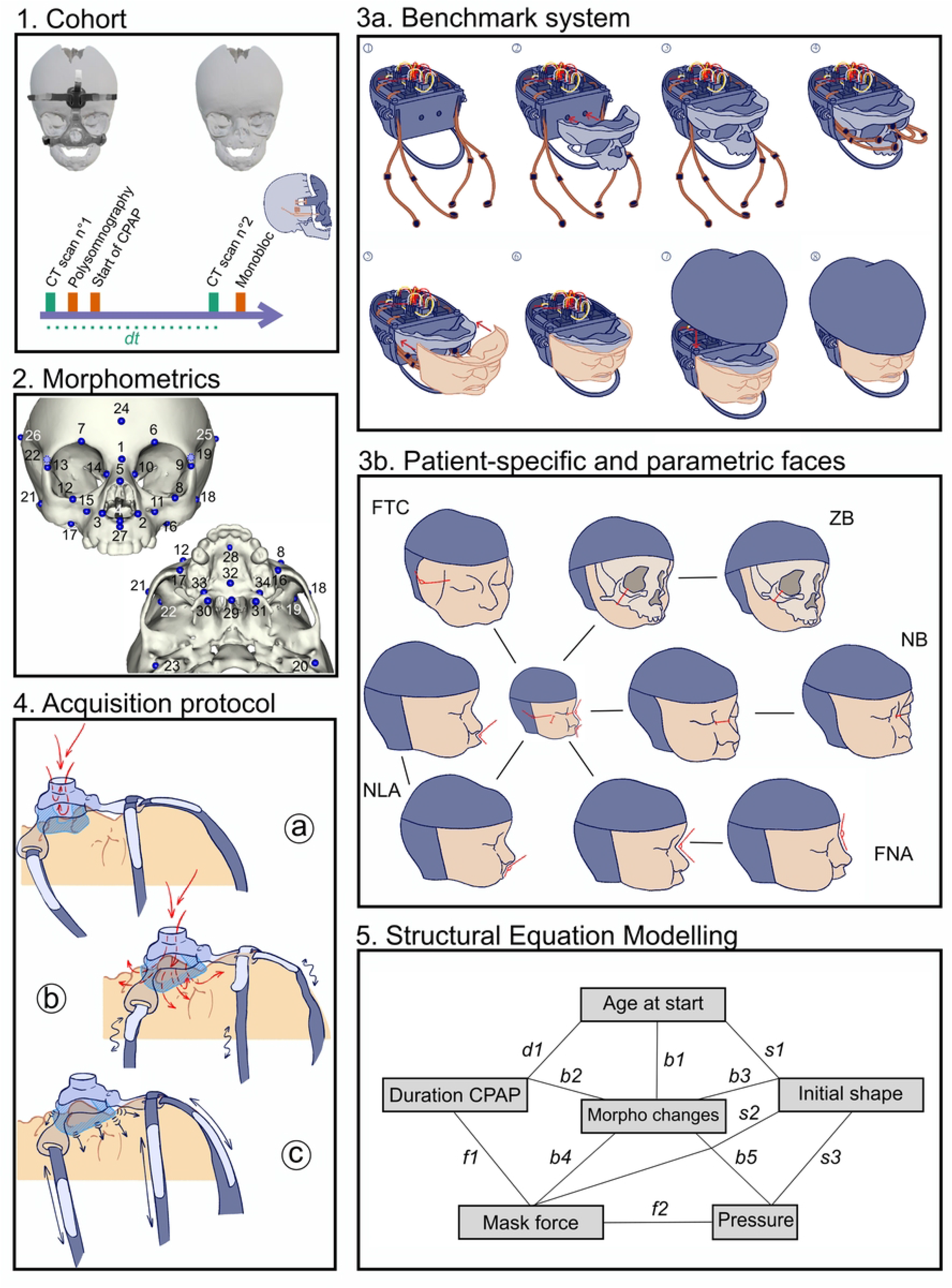
Pipeline of data acquisition and analyses. (2.1) Management and imaging schedule of the cohort. (2.2) Thirty-four landmarks manually positioned on CT scans (details in S2 Table). (2.3a) Benchmark design: fitting of five 3D-printed modules: (1) electronics housing module, (2) skull base module, (3) midfacial bony module, (4) cranial vault module, and (5) midfacial soft-tissue module. (2.3b) Ten parametric facial modules based on two extreme values of five parameters: (1) fronto-temporal curvature (FTC), (2) zygoma breadth (ZB), (3) frontonasal angle (FNA), (4) nasolabial angle (NLA), and (5) nasal bridge ratio (NB). (2.4) Acquisition protocol using the CPAP device: (a) positioning the mask on the nose, (b) adjustment of the mask to minimize unintentional air leaks, and (c) adapted mask tightening. (2.5) Structural equation modeling: morphological changes between CT1 and CT2 are considered to be driven by five main factors, the links between variables expressing correlation hypotheses.

#### Experimental assessment of mask-induced mechanical loading

To quantify mask–face mechanical interactions, we developed a modular system combining patient-specific and parametric 3D-printed facial models with embedded pressure sensors (Fig 2.3, S1–S7 Figs, S3 and S4 Tables). Contact pressure was recorded at four midfacial sites using capacitive sensors, and overall mask tightening force with a dynamometer. Repeated measurements were obtained with a standardized nasal mask at 8 cmH_2_O across operators, sessions, and fitting configurations (minimal leak versus intentional overtightening) (Fig 2.4, S8 Fig).

#### Structural equation modeling

Structural equation modeling (SEM) was used to test a predefined causal framework (Fig 2.5) linking craniofacial shape change with age at CT1, initial craniofacial morphology, duration of CPAP use, and mechanical loading (force and pressure) [25].

Initial morphology was represented by the first principal component of CT1 shape (PC1_shape_), while craniofacial outcome was quantified as the magnitude of shape change between CT1 and CT2 (displacement vectors). Mechanical exposure was described by mask tightening force (provided by the dynamometer) and pressure distribution (principal component of sensor measurements). Age at CT1 and duration of CPAP use were included as metadata. Path coefficients were estimated using maximum likelihood, and standard errors were obtained by bootstrap resampling (2000 iterations). Model fit was assessed using the χ² test and the Standardized Root Mean Square Residual (SRMR). Indirect effects were computed as the product of path coefficients along predefined pathways using the *lavaan* package in R.

## Results

### Baseline morphology

Pairwise comparisons using Procrustes ANOVA showed that baseline morphology differed between the mask group and controls irrespective of age (R² = 0.237, F = 11.269, Z = 3.920, p = 0.001), and likewise between the no-mask group and controls (R² = 0.166, F = 5.924, Z = 3.716, p = 0.002). By contrast, the mask and no-mask groups did not differ morphologically at CT1, with or without accounting for age (R² = 0.043, F = 1.263, Z = 0.760, p = 0.230; R² = 0.028, F = 0.831, Z = −0.176, p = 0.577) (Fig 3.1). This established a comparable baseline for the longitudinal analyses and indicated that subsequent divergence could not be attributed to a major initial shape imbalance alone.

**Fig 3.**
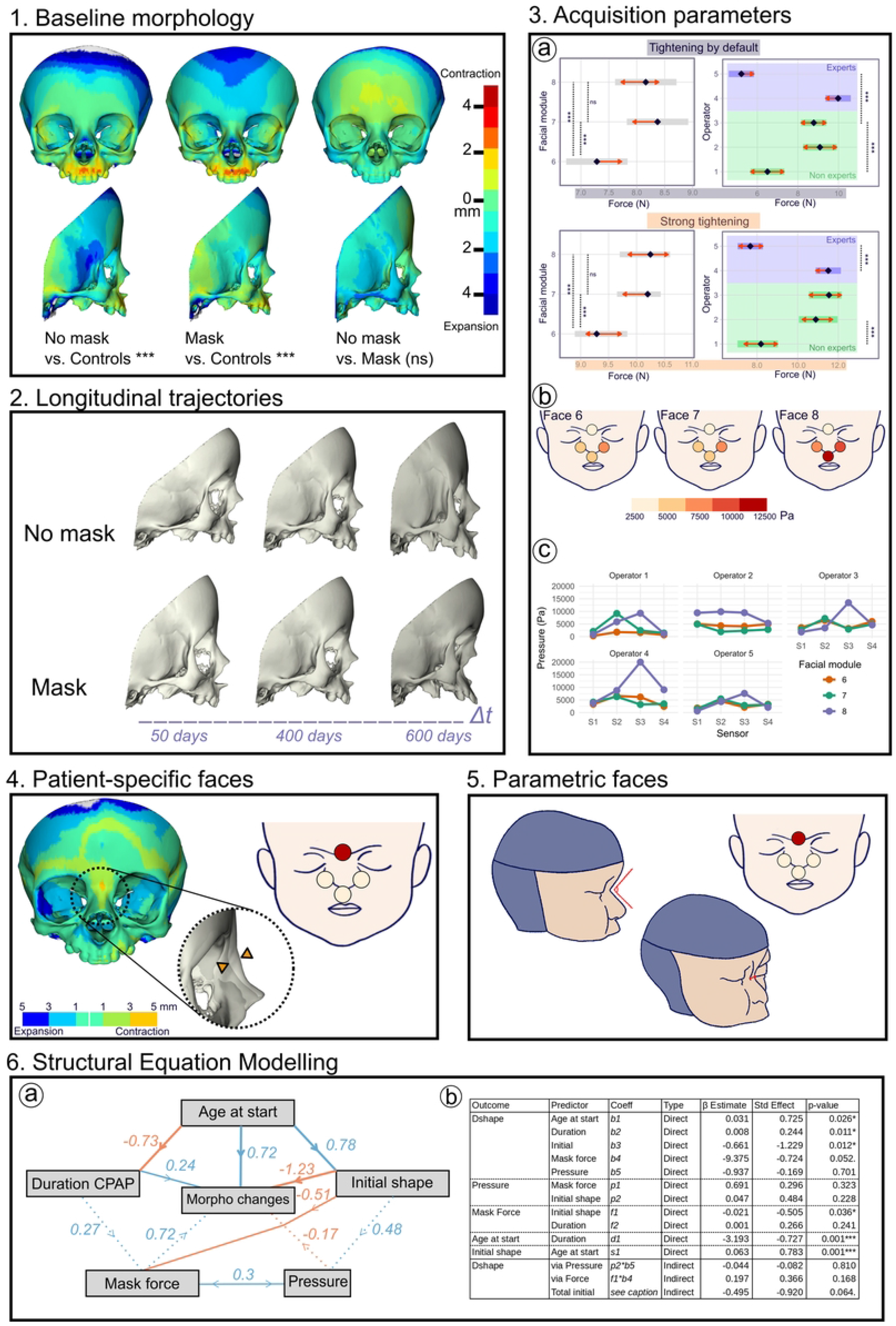
Results. (3.1) Pairwise comparisons of baseline morphology between groups (*** p < 0.001; ns, not significantly different). (3.2) Longitudinal trajectories of the no-mask and mask groups between CT1 and CT2 (Δt, in days): pronounced midfacial retrusion in the mask group compared with typical growth impairment in the no-mask group. (3.3a) Effect of operator and facial modules on mask tightening force (N), with normal tightening and with overtightening: increase of absolute force but preserved effects of operator and facial shape. (3.3b) Mean pressure distribution (Pa) across sensors in the three faces tested. (3.3c) Mean pressure distribution across sensors according to each operator (S1, nasion; S2 and S3, nasal alae; S4, anterior maxilla between subnasale and prosthion). (3.4) Main axis of covariation between patient-specific initial facial shape and measured pressure distribution: retruded nasal root associated with higher pressure concentration captured by sensor S1 located on the nasal root. (3.5) Correlation between nasal root and frontal bossing with measured mask tightening and pressure distribution. (3.6) Structural equation modeling. (3.6a) Significant (solid lines) and non-significant (dashed lines) standardized correlations (positive in blue, negative in orange); line thickness proportional to the standardized beta estimate (Std Effect). (3.6b) All direct and indirect paths tested to explain shape deformation (Dshape). The predictor “total initial” tested b3 + (f1 × b4) + (p2 × b5) + (f1 × p1 × b5), that is, the full mechanical cascade from the initial experimental conditions potentially driving Dshape.

### Risk factor 1: age at treatment initiation

Age at treatment initiation was associated with subsequent craniofacial deviation: younger treated patients followed a more retrusive developmental trajectory than those starting later. In patient-specific models, younger patients with smaller faces were also associated with higher tightening forces (Fig 3.2, 3.4 and 3.6a).

### Risk factor 2: duration of CPAP treatment

The effect of Δt on longitudinal shape change was significant in both groups, but markedly stronger in the mask group (no-mask: r_PLS_ = 0.720, p = 0.001; mask: r_PLS_ = 0.865, p = 0.001). In the no-mask group, growth patterns were characterized by facial flattening with zygomatic retrusion and vertical forehead elongation. In the mask group, by contrast, shape change was characterized by pronounced retrusion of the midface, associated with counter-clockwise rotation of the maxilla, clockwise rotation of the nasal region, and frontal bossing. These changes were visible after 1 year of CPAP use and became more obvious after 2 years (Fig 3.2), supporting a cumulative exposure effect distinct from untreated growth.

### Risk factor 3: initial morphology

Initial morphology predicted both mechanical exposure and craniofacial outcome. CT1 PC1 significantly predicted applied force (p < 0.01), while contact pressure distribution varied with morphology, particularly nasal root retrusion (Fig 3.4). Similar relationships were observed in parametric models (Fig 3.5). Nasal root retrusion with frontal bossing was associated with higher forces and more localized pressure concentration, defining a clinically relevant high-risk morphology. Contact pressure alone did not predict subsequent shape change.

### Associated mask-induced force and pressure patterns

CPAP masks applied substantial forces to the patient-specific facial models, with tightening levels ranging from 6 to 10 N (Fig 3.3a). Force measurements were repeatable across sessions. A significant operator effect was observed (F = 38.0, p < 0.001), although individual operators were consistent in their tightening behavior. Facial geometry also significantly influenced applied force (F = 4.91, p = 0.011), with specific facial configurations consistently associated with higher tightening levels (Fig 3.3a). When comparing fitting conditions, intentional overtightening resulted in a marked increase in applied force (mean difference ≈ 4.06 N; F = 276.8, p < 0.001) (Fig 3.3a). Notably, no clear difference was observed between experienced operators and non-expert users in the magnitude of applied force.

Contact pressure measurements revealed structured spatial patterns across sensors (F = 11.83, p < 0.001), with a strong interaction between sensor location and facial geometry (F = 12.44, p < 0.001), indicating that pressure distribution depended primarily on craniofacial morphology. In contrast, operator effects on contact pressure distribution were limited (sensor × operator interaction: F = 1.85, p = 0.042) (Fig 3.3c).

### Integrated model linking age, duration, morphology, loading, and shape change

A causal framework linking craniofacial shape change with age at CPAP initiation, initial morphology, duration of CPAP use, and mechanical loading was tested using SEM (Fig 3.6a). Although model fit was only moderate (χ² = 3.17, df = 4, p = 0.53; SRMR = 0.087), it explained a substantial proportion of variance in shape change (R² = 0.67, p = 0.006). Initial craniofacial morphology was the dominant determinant of shape change (standardized effect β = −1.23, p = 0.012), while duration of CPAP use contributed to a lesser extent (β = 0.24, p = 0.011). Force showed a borderline association with shape change (β = −0.72, p = 0.052), whereas pressure distribution was not predictive (β = −0.17, p = 0.701). Initial morphology also significantly influenced mechanical exposure, predicting applied force (β = −0.51, p = 0.036) and, to a lesser extent, pressure distribution. Indirect pathways linking morphology to outcome through force and pressure were not significant (Fig 3.6b).

## Discussion

Our longitudinal and experimental data indicate that prolonged nasal CPAP is not mechanically neutral during craniofacial growth and is associated with additional midfacial retrusion beyond the natural course of *FGFR*-related craniosynostoses.

### Clinical risk factors and biomechanical interpretation

Our findings support a biomechanical interpretation in which CPAP-related change is not an immediate deformation proportional to a single measured force or pressure value, but rather a growth deviation developing under sustained constraint. Craniofacial growth involves coordinated remodeling of bone, sutures, cartilage, and soft tissues [13], and orthodontic or orthopedic treatments provide well-known examples of prolonged extraoral forces redirecting facial growth [12,26]. Force magnitude and local pressure distribution accounted for the magnitude of deformation but not directly for specific deformation patterns. Rather, age at onset, initial morphology, and duration appeared to define the developmental context within which mechanical loading was expressed, consistent with a non-linear growth response under sustained constraint. The absence of a direct relationship between local contact pressure and shape change pattern does not imply that pressure is irrelevant; rather, it likely reflects the fact that real-life mask position, orientation, and fitting may vary over days and months, whereas our controlled measurements capture only standardized snapshots of this mechanical environment.

### Mask-induced force and pressure patterns

Our experimental results are in line with previous studies showing that CPAP nasal masks generate heterogeneous pressure patterns concentrated on anatomically prominent regions, especially the nasal root and upper lip region [18,19]. A more retruded nasal root, combined with frontal bossing, was associated with higher tightening forces and greater pressure concentration at the nasal root, likely because these morphologies make it more difficult to obtain an airtight seal with standard interfaces. This interpretation is consistent with previous reports that mask fit and treatment performance depend on craniofacial phenotype, and that a given interface may behave differently across facial morphologies [18,20].

Although tightening force differed significantly between operators, no clear difference was observed between expert and non-expert users. This suggests that mechanical loading is driven less by expertise than by anatomy and fitting constraints. This is clinically important because it implies that unfavorable loading may persist even when masks are fitted carefully and consistently.

### Implications for craniofacial management

The risk associated with CPAP use is unlikely to be uniform across patients. Our results suggest that nasal root retrusion and frontal bossing are potentially important clinical markers of vulnerability to unfavorable mask loading. This opens the possibility of identifying, before or at CPAP initiation, patients at higher mechanical risk with standard nasal masks. Such patients may require closer longitudinal monitoring, earlier reassessment of mask fit, alternative interface strategies, or broader discussion of treatment sequencing. Identification of such higher-risk morphologies may therefore inform longitudinal craniofacial surveillance and multidisciplinary discussions regarding the timing of respiratory and surgical interventions. Timing and CPAP duration are also central. The use of prolonged CPAP during early childhood may amplify growth deviation under constraint. Clinically, this supports systematic follow-up of craniofacial growth in treated patients and repeated multidisciplinary reassessment of mask fit, treatment duration, and the balance between CPAP, upper airway surgery, tracheostomy, and craniofacial advancement. Current pediatric interfaces are not designed for the craniofacial anatomy of children with syndromic craniosynostoses, despite growing recognition that mask selection and interface personalization are determinants of efficacy and tolerance [20]. Morphology-adapted masks, including designs accommodating nasal inclination and redistributing pressure away from vulnerable areas such as the nasal root, may reduce localized loading and improve safety [27,28]. This is especially relevant for the management of patients with achondroplasia, an *FGFR3*-related condition with a high prevalence of OSA and a craniofacial phenotype characterized by midfacial hypoplasia, frontal bossing, and a depressed nasal root [29].

### Limitations and future directions

Our sample size was limited, particularly for SEM, and the model therefore requires validation in larger cohorts. The no-mask group also introduces indication bias: absence of CPAP reflected lower respiratory severity, which may capture clinical differences not fully represented by craniofacial shape. Although baseline morphology did not differ detectably between groups after accounting for age, residual heterogeneity within and across *FGFR*-related syndromes remains possible [30].

Only one mask was tested experimentally, we did not compare the Nonny interface, and daily hours of CPAP use were unavailable despite their potential contribution to cumulative exposure. Sensitivity analyses across CPAP levels (6–12 cmH_2_O) indicated that pressure level did not influence total force or pressure patterns (S1 Text). Finally, we cannot yet determine whether the observed growth deviation translates into differences in surgical trajectory, surgical difficulty, or long-term respiratory outcome.

Although these growth-related findings are specific to children with *FGFR*-related craniosynostoses, the methodological framework is not disease-specific. Integrating longitudinal morphology with mask tightening force and pressure distribution could be applied to other pediatric or adult populations to characterize patient–interface interactions and support more individualized interface selection.

Future work should validate these findings in larger multicentric cohorts, including through the European Reference Network ERN CRANIO, and integrate age at initiation, treatment duration, baseline morphology, daily use, and genotype. Prospective studies comparing multiple interfaces should also determine whether reducing mask-induced loading improves preservation of craniofacial growth and subsequent management.

## Conclusions

CPAP use in infants with *FGFR*-related craniosynostoses is associated with additional midfacial growth deviation and should not be considered mechanically neutral during craniofacial development. Younger age at initiation, longer CPAP use, and specific baseline morphologies, especially nasal root retrusion with frontal bossing, appear to define higher-risk situations. These findings support systematic craniofacial growth monitoring during prolonged CPAP and consideration of respiratory treatment exposure within multidisciplinary craniofacial management. They also support the development of morphology-adapted interfaces in children requiring prolonged CPAP. Further work is now needed to determine whether these early growth effects influence long-term craniofacial outcome, respiratory trajectory, and later facial surgical management.

## Data Availability

The minimal data set and accompanying code are available on the website of the Craniofacial Growth and Form laboratory (growthnform.org).

https://growthnform.org

## Acknowledgments

We are grateful to Marine Dosso (nurse in the NIV and sleep unit at Necker – Enfants malades Hospital) for her participation in the experimental part on the evaluation of mask contact pressures.

## Supporting information

**S1 Checklist. STROBE checklist.** Completed STROBE checklist for the observational component of the study.

**S1 Text. Supplementary methods.** Design of the modular 3D-printed benchmark (electronics housing, skull base, midfacial bony, cranial vault and midfacial soft-tissue modules), Arduino architecture and acquisition code, definition of the morphometric parameters, segmentation and CAD workflow of the patient-specific models, construction of the parametric models, 3D printing and materials, sensor system, force and pressure acquisition protocol, and architecture of the structural equation model.

**S1 Table. Characteristics of the patients and controls.** Syndrome and genotype, age at CPAP start, at CT1, at ENT and posterior cranial vault surgery, at CT2 and at CPAP stop, CPAP duration and level, and interface, for the mask group, the no-mask group and the healthy controls.

**S2 Table. Landmark definitions.** Definitions of the 34 landmarks placed on the midface and forehead.

**S3 Table. Morphometric parameters across the cohort.** Mean, standard deviation and coefficient of variation of the five morphometric parameters (zygoma length, frontonasal angle, nasolabial angle, zygomatico-temporal angle, nasal bridge ratio).

**S4 Table. Extreme values of the five parameters.** Extreme values used to print the parametric head models.

**S1 Fig. General design of the 3D-printed benchmark.** Fixed module holding the Arduino system, rigid support holding the skeletal module, silicone soft-tissue module and rigid skull vault module.

**S2 Fig. Arduino setup.** Arduino UNO Rev3 architecture and connection of the four pressure sensors.

**S3 Fig. Segmentation of the patient-specific bony parts and soft tissues.** (A) Bone mask on a sagittal CT slice. (B) Soft-tissue mask and 3D reconstruction.

**S4 Fig. Morphometric parameters.** (A) Anatomical landmarks and angles on the 3D soft-tissue isosurface. (B) Landmarks and distances with bones in transparency, showing the zygoma and the zygomaxillare point. (C) The five parameters measurable by direct clinical evaluation.

**S5 Fig. Parametric mesh deformation.** Anatomical vertices selected in Blender on the soft-tissue and bone meshes for the five parameters, and controlled translation of these vertices until the target value was reached.

**S6 Fig. The ten 3D-printed parametric head models used for pressure measurements.**

**S7 Fig. Quasi-static compression tests.** Instron compression tests on a single sensor under different silicone thicknesses: test plates, experimental setup, and applied versus measured pressure.

**S8 Fig. Final sensor system and complete benchmark setup.** Sensors plugged into the midfacial bony module, and the assembled benchmark with the nasal mask and CPAP device.

